# SARS-CoV-2 Seroprevalence Among Firefighters in Los Angeles, California

**DOI:** 10.1101/2021.06.03.21258299

**Authors:** Karen Mulligan, Anders H. Berg, Marc Eckstein, Acacia Hori, Anna Rodriguez, Omar Toubat, Neeraj Sood

## Abstract

**Objective:** We estimate the seroprevalence of SARS-CoV-2 antibodies among firefighters in the Los Angeles, California fire department in October 2020 and compare demographic and contextual factors for seropositivity.

**Methods:** We conducted a serologic survey of firefighters in Los Angeles, California, USA, in October 2020. Individuals were classified as seropositive for SARS-CoV-2 if they tested positive for immunoglobulin G, immunoglobulin M, or both. We compared demographic and contextual factors for seropositivity.

**Results:** Of 713 participants, 8.9% tested positive for SARS-CoV-2 antibodies. Seropositivity was not associated with gender, age, or race/ethnicity. Furthermore, firefighters who worked in zip codes with lower income or higher share of minority population did not have higher rates of SARS-CoV-2 infection. Seropositivity was highest among firefighters who reported working in the vicinity of Los Angeles International Airport, which had a known outbreak in July 2020.

**Conclusions:** Seroprevalence among firefighters was no higher than seroprevalence in the general population, suggesting that workplace safety protocols, such as access to PPE and testing, can mitigate increased risk of infection at work. Workplace safety protocols for firefighters also eliminated differences in disease burden by geography and race/ethnicity observed in the general population.

## INTRODUCTION

First responders–including firefighters–have been working throughout the pandemic, which increases the likelihood of contracting severe acute respiratory syndrome coronavirus 2 (SARS-CoV-2). While we may expect higher exposure to lead to higher rates of infectivity among first responders relative to the general population, preventive measures such as using personal protective equipment (PPE), conducting workplace screening, or implementing protocols to minimize exposure to infected patients may mitigate the increased risk of contracting SARS-CoV-2 among first responders. Measuring seroprevalence of SARS-CoV-2 among first responders will improve our understanding of risk and transmission in front line worker populations.

Several studies have estimated seroprevalence in firefighter populations during the first six months of the pandemic in 2020. On the lower end, seroprevalence estimates ranged from 1.1% and 1.5% in Rochester (May) and Arizona (April/May), respectively.[1,2] In May/June seroprevalence was 5.4% in Cleveland 6.9% in Detroit.[3,4] In a single fire department located at the epicenter of an outbreak in South Florida, seroprevalence was 8.9% in April.[5] Across all published studies, seroprevalence was highest among New York City firefighters: 22.5% in May-July.[6] Although it has been demonstrated that healthcare workers with high levels of patient contact experience greater odds of seropositivity than the general population or other public service employees,[4,7] seroprevalence among first responders or firefighters do not appear to significantly deviate from community estimates.[1,2,6]

In this study we conducted a serologic survey to estimate the prevalence of SARS-CoV-2 antibodies among firefighters in the Los Angeles Fire Department (LAFD) and tested for associations between seroprevalence and individual characteristics.

## METHODS

### Study design

This is a prospective cohort study of firefighters employed by LAFD conducted in partnership between the University of Southern California, the LA County Department of Public Health, LAFD, Gauss Surgical, and Cedars Sinai Medical Center. Firefighters were invited to complete a questionnaire and received polymerase chain reaction (PCR) and antibody tests. This study was approved by the Los Angeles (LA) County Department of Public Health Institutional Review Board. Written informed consent was obtained from all study participants.

### Study participants and data

An estimated 3,000 firefighters actively employed by LAFD were eligible for study participation. Participants were recruited through an employee intranet between July and October 2020. Participant onboarding was performed through a proprietary web- and mobile-based application developed by Gauss Surgical (Menlo Park, CA.) Onboarding involved describing study aims and methods to the participant, consenting the participant to complete PCR and antibody tests, and scheduling test dates. Data on participant age, sex, race/ethnicity, workplace location, symptoms, hospitalizations, patterns of PPE use, and PCR-confirmed SARS-CoV-2 infection history were collected through electronic surveys (Qualtrics International Inc., Seattle, WA).

### PCR and antibody testing

PCR and antibody testing occurred between October 20 and 28, 2020. Participants who self-reported symptoms on a COVID-19 symptom screener on the test day were excluded from the study (N=4). PCR tests were performed on oropharyngeal specimens self-collected by participants under study staff observation. RT-PCR was conducted against E and S mRNA transcripts at Cedars-Sinai Medical Center Department of Pathology (Los Angeles, CA.) For seroprevalence assessments, certified phlebotomists collected approximately 5 milliliters of venous blood from each participant. SARS-CoV-2-specific immunoglobulin M (IgM) and immunoglobulin G (IgG) antibodies were measured using the Abbott Architect instrument (Abbott Laboratories, Chicago, IL.) Manufacturer recommended signal-to-cutoff (S/CO) ratios of >1.4 and >1.0 were used as thresholds for seropositivity for IgG and IgM antibodies, respectively.

### Statistical analysis

Participants were defined as seropositive if they had IgG, IgM, or both types of antibodies. Descriptive statistics for survey responses were summarized for the study cohort. We tested for differences in seropositivity by participant characteristics using two-sided tests and a significance level of p<0.05. We also performed secondary analyses, including multivariable logistic regression and testing for differences in firefighter seroprevalence by contextual characteristics of workplace zip codes. We conducted sensitivity analysis using a modified S/CO threshold based on results from a validation study.[8] Results for the secondary and sensitivity analyses are presented in the Appendix. All statistical analyses were performed using Stata (StataCorp LLC, College Station, TX).

## RESULTS

713 out of 715 firefighters who participated in the study had valid serologic data. Survey respondents had similar demographics to the overall population of LAFD. We limited our analysis sample to the 686 participants with no missing values for survey questions. Among the firefighters that participated in the study and had non-missing data for survey questions, 61 (8.9%) tested positive for either IgG, IgM, or both types of antibodies (Table 1). Approximately 14% of our respondents reported that they have not received a previous PCR test and 7.0% of respondents who reported not receiving a PCR test were antibody positive. Additional undiagnosed cases occurred among those who received a previous PCR test and got a negative result, with estimated seroprevalence of 3.2%.

**Table 1.**
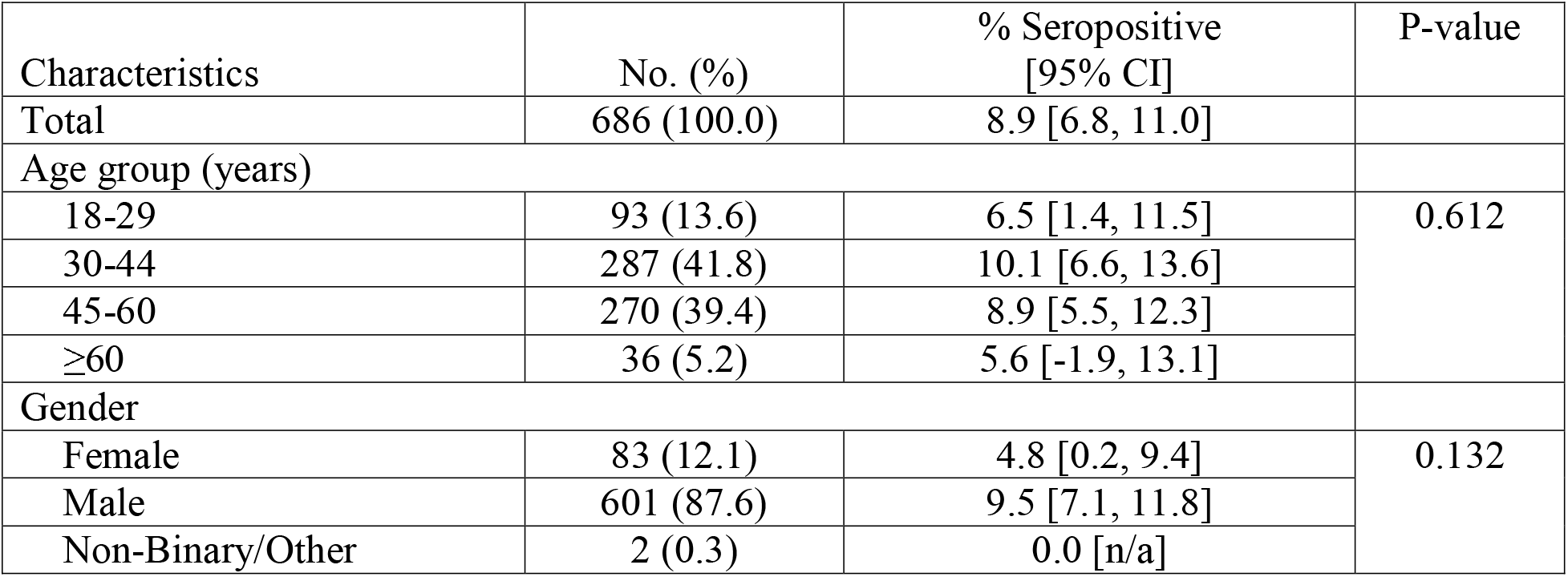
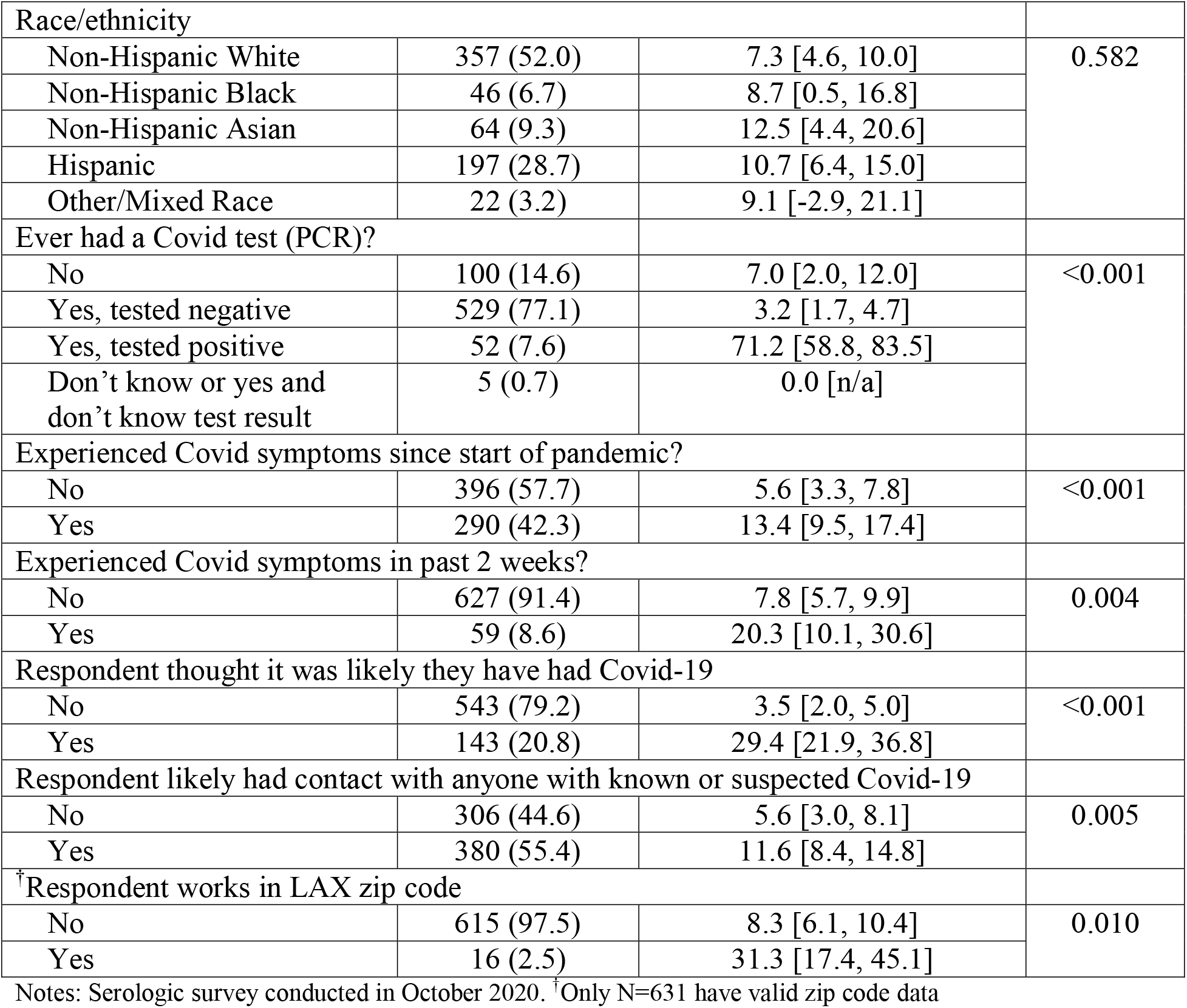
SARS-CoV-2 seropositivity and association with individual characteristics among firefighters in Los Angeles, CA, USA.

| Characteristics | No. (%) | % Seropositive [95% CI] | P-value |
| --- | --- | --- | --- |
| Total | 686 (100.0) | 8.9 [6.8, 11.0] |  |
| Age group (years) |  |  |  |
| 18-29 | 93 (13.6) | 6.5 [1.4, 11.5] | 0.612 |
| 30-44 | 287 (41.8) | 10.1 [6.6, 13.6] |  |
| 45-60 | 270 (39.4) | 8.9 [5.5, 12.3] |  |
| ≥60 | 36 (5.2) | 5.6 [-1.9, 13.1] |  |
| Gender |  |  |  |
| Female | 83 (12.1) | 4.8 [0.2, 9.4] | 0.132 |
| Male | 601 (87.6) | 9.5 [7.1, 11.8] |  |
| Non-Binary/Other | 2 (0.3) | 0.0 [n/a] |  |
| Race/ethnicity |  |  |  |
| Non-Hispanic White | 357 (52.0) | 7.3 [4.6, 10.0] | 0.582 |
| Non-Hispanic Black | 46 (6.7) | 8.7 [0.5, 16.8] |  |
| Non-Hispanic Asian | 64 (9.3) | 12.5 [4.4, 20.6] |  |
| Hispanic | 197 (28.7) | 10.7 [6.4, 15.0] |  |
| Other/Mixed Race | 22 (3.2) | 9.1 [-2.9, 21.1] |  |
| Ever had a Covid test (PCR)? |  |  |  |
| No | 100 (14.6) | 7.0 [2.0, 12.0] | <0.001 |
| Yes, tested negative | 529 (77.1) | 3.2 [1.7, 4.7] |  |
| Yes, tested positive | 52 (7.6) | 71.2 [58.8, 83.5] |  |
| Don't know or yes and don't know test result | 5 (0.7) | 0.0 [n/a] |  |
| Experienced Covid symptoms since start of pandemic? |  |  |  |
| No | 396 (57.7) | 5.6 [3.3, 7.8] | <0.001 |
| Yes | 290 (42.3) | 13.4 [9.5, 17.4] |  |
| Experienced Covid symptoms in past 2 weeks? |  |  |  |
| No | 627 (91.4) | 7.8 [5.7, 9.9] | 0.004 |
| Yes | 59 (8.6) | 20.3 [10.1, 30.6] |  |
| Respondent thought it was likely they have had Covid-19 |  |  |  |
| No | 543 (79.2) | 3.5 [2.0, 5.0] | <0.001 |
| Yes | 143 (20.8) | 29.4 [21.9, 36.8] |  |
| Respondent likely had contact with anyone with known or suspected Covid-19 |  |  |  |
| No | 306 (44.6) | 5.6 [3.0, 8.1] | 0.005 |
| Yes | 380 (55.4) | 11.6 [8.4, 14.8] |  |
| † Respondent works in LAX zip code |  |  |  |
| No | 615 (97.5) | 8.3 [6.1, 10.4] | 0.010 |
| Yes | 16 (2.5) | 31.3 [17.4, 45.1] |  |

Notes: Serologic survey conducted in October 2020. † Only N=631 have valid zip code data

Gender, race, and age were not statistically significant predictors of seropositivity. Seropositivity was lower among females (odds ratio (OR)=0.48, 95% CI=0.17-1.3), but the difference between men and women was not statistically significant. Seroprevalence was not statistically different across race or ethnicity, but point estimates were highest among non-Hispanic Asian participants (OR=1.82, 95% CI=0.78-4.2). We found no statistical differences in seroprevalence by age, but the point estimate was lowest among firefighters aged over 60 years (OR=0.85, 95% CI=0.16-4.42). Seroprevalence was highest (31.3% (OR=4.98, 95% CI=1.67-14.9) among participants who reported working in the zip code containing the Los Angeles International Airport (LAX).

## DISCUSSION

SARS-CoV-2 seroprevalence was 8.9% in LAFD in October 2020. This estimate is 1 to 3 percentage points higher compared with estimates from other firefighter populations taken earlier in the pandemic (excluding NYC.) Cumulative incidence in our sample could be as high as 11.1% if we include participants who were not seropositive but received a positive PCR test (i.e., those who likely had SARS-CoV-2 but no longer have antibodies) in our calculations. While firefighters have higher risk of contracting SARS-CoV-2 due to the nature of their work, seroprevalence in our sample is comparable to that in the general LA population during the same timeframe (15%.)[9,10]

We found no evidence that firefighter demographics were associated with increased risk of seropositivity. The relatively small sample size in our study and corresponding low power for statistical tests provides one possible explanation for this result. Alternatively, lack of demographic differences in seropositivity may reflect uniform implementation of SARS-CoV-2 workplace protocols by LAFD such as wearing N95 masks, goggles, and gloves for all EMS incidents as well as putting masks on patients at time of first contact.[11] Analysis of contextual factors potentially supports this explanation: firefighters who work in zip codes that were disproportionately impacted by SARS-CoV-2 (i.e.,those with lower median income or a higher share of non-white, non-Hispanic residents[12]) did not have higher seroprevalence rates.

LAFD firefighters–including those who are asymptomatic–have had open access to PCR testing, which includes four sites located at fire stations throughout LA. Despite this, approximately 14% of our respondents (including 10% of those who reported experiencing symptoms during the pandemic) reported that they have not received a PCR test. Consequently, potential SARS-CoV-2 cases went undiagnosed: 7.0% of respondents who reported not receiving a PCR test were antibody positive. Additional undiagnosed cases occurred among those who received PCR tests and got a negative result, with estimated seroprevalence of 3.2%. This suggests that even workplaces that have relatively easy access to PCR testing may benefit from more systematic testing to identify asymptomatic or mildly symptomatic infections and limit workplace exposure.

This work should be viewed in light of its limitations. Our sample was self-selected, and accounted for approximately 24% of the full LAFD employee population. Participation may have been influenced by prior testing results, household exposure, or worker availability. Nevertheless, our sample demographics were representative of the overall LAFD population. Second, we did not ask respondents questions about their home environment–such as the total number of residents or the size of their home–which could have influenced their exposure to SARS-CoV-2. Furthermore, our sample collection period occurred in the months following the first wave of the pandemic, yet prior to the substantial rise in SARS-CoV-2 cases in LA County and the US beginning in November 2020.[13] Despite this, our study provides seroprevalence estimates and factors associated with SARS-CoV-2 infection for a population that is both at high risk of coming in contact with SARS-CoV-2, but also follows strong workplace protection practices.

Between 8.9% and 11.0% of firefighters in LAFD were infected with SARS-CoV-2 depending on whether we adjustment for waning antibodies. We did not observe significant differences in seroprevalence by demographic factors. Furthermore, our results suggest workers in occupations that adhere to similar protection or mitigation workplace protocols are likely to experience similar rates of SARS-CoV-2 infection irrespective of workplace location.

## Data Availability

In order to maintain privacy of study participants, data cannot be shared publicly.

## Acknowledgements

None

