## Appendix for "SARS-CoV-2 Seroprevalence Among Firefighters in Los Angeles, California"

**Table 1. SARS-CoV-2 seroprevalence among firefighters in Los Angeles, California, USA, October 2020**

| Sample | Sample size | Number (%) Seropositive | Number reported having a past PCR test (% with positive result) |
| --- | --- | --- | --- |
| Valid serologic data | 713 | 63 (8.8) | 607 (8.9) |
| Non-missing survey data | 686 | 61 (8.9) | 586 (8.9) |
| Valid zip code data | 631 | 56 (8.9) | 540 (9.1) |

Notes: Individuals were classified as being seropositive if they had IgG, IgM, or both types of SAR-CoV-2 antibodies.

Our main analysis used signal-to-cutoff (S/CO) thresholds specified by the manufacturer. Because these thresholds were developed to detect antibodies in the weeks following hospitalization for SARS-CoV-2 infection when titers are highest and recent studies have shown that SARS-CoV-2 antibody titers are lower for those with mild disease and often decrease within months following recovery,[1,2] this threshold may be too high to sensitively detect antibodies months after recovery. For this reason, we performed validation studies to determine whether a lower S/CO threshold could be used to more sensitively detect antibodies while still remaining diagnostically specific for SARS-CoV-2 infection. SARS-CoV-2 IgG and IgM antibody titers were measured in 178 negative control patient blood samples collected before the beginning of the pandemic (before January 2020).[3] Our analysis determined that S/CO ratio values of 0.36 for IgG titers and 0.62 for IgM titers were optimal and even at these lower cut-offs test specificity was 99.4%. These values were used as a modified threshold for seropositivity.

**Table 2. Comparison of Results for Manufacturer and Modified Signal-to-Cutoff Values**

| Characteristics | No. (%) | % Seropositive [95% CI] | |
| --- | --- | --- | --- |
|  |  | Manufacturer cutoff | Modified cutoff |
| Total | 686 (100.0) | 8.9 [6.8, 11.0] | 13.4 [11.0, 16.1] |
| Age group (years) | | | |
| 18-29 | 93 (13.6) | 6.5 [1.4, 11.5] | 12.9 [6.9, 21.0] |
| 30-44 | 287 (41.8) | 10.1 [6.6, 13.6] | 15.0 [10.9, 19.1] |
| 45-60 | 270 (39.4) | 8.9 [5.5, 12.3] | 13.0 [9.0, 17.0] |
| ≥60 | 36 (5.2) | 5.6 [-1.9, 13.1] | 5.6 [-1.9, 13.0] |
| Gender | | | |
| Female | 83 (12.1) | 4.8 [0.2, 9.4] | 8.4 [1.1, 15.8] |
| Male | 601 (87.6) | 9.5 [7.1, 11.8] | 14.1 [11.5, 17.1] |
| Non-Binary/Other | 2 (0.3) | 0.0 [n/a] | 0.0 [n/a] |
| Race/ethnicity | | | |
| Non-Hispanic White | 357 (52.0) | 7.3 [4.6, 10.0] | 12.3 [8.9, 15.7] |
| Non-Hispanic Black | 46 (6.7) | 8.7 [0.5, 16.8] | 15.2 [6.4, 28.3] |
| Non-Hispanic Asian | 64 (9.3) | 12.5 [4.4, 20.6] | 12.5 [4.4, 20.6] |
| Hispanic | 197 (28.7) | 10.7 [6.4, 15.0] | 14.2 [9.3, 19.1] |
| Other/Mixed Race | 22 (3.2) | 9.1 [-2.9, 21.1] | 22.7 [5.2, 40.2] |
| Ever had a Covid test (PCR)? |  |  |  |
| No | 98 (14.3) | 6.1 [1.4, 10.9] | 9.2 [3.5, 14.9] |
| Yes | 586 (85.4) | 9.2 [6.9, 11.6] | 14.0 [11.3, 17.0] |
| Don’t know | 2 (0.3) | 50.0 [n/a] | 50.0 [n/a] |
| Covid test result ^x,y^ | | | |
| Negative | 529 (90.3) | 3.2 [1.7, 4.7] | 6.6 [4.6, 9.0] |
| Positive | 52 (8.9) | 71.2 [58.8, 83.5] | 90.4 [83.4, 97.3] |
| Don’t know | 5 (0.9) | 0.0 [n/a] | 0.0 [n/a] |
| Experienced Covid symptoms since start of pandemic? ^y^ | | | |
| No | 396 (57.7) | 5.6 [3.3, 7.8] | 9.3 [6.2, 12.9] |
| Yes | 290 (42.3) | 13.4 [9.5, 17.4] | 19.0 [15.0, 22.9] |
| Experienced Covid symptoms in past 2 weeks? | | | |
| No | 627 (91.4) | 7.8 [5.7, 9.9] | 12.8 [10.3, 15.5] |
| Yes | 59 (8.6) | 20.3 [10.1, 30.6] | 20.3 [10.1, 30.6] |
| Respondent thought it was likely they have had Covid-19 ^x,y^ | | | |
| No | 543 (79.2) | 3.5 [2.0, 5.0] | 6.6 [4.0, 9.3] |
| Yes | 143 (20.8) | 29.4 [21.9, 36.8] | 39.9 [34.7, 45.0] |
| Respondent likely had contact with anyone with known or suspected Covid-19 ^x,y^ | | | |
| No | 306 (44.6) | 5.6 [3.0, 8.1] | 8.8 [5.0, 12.6] |
| Yes | 380 (55.4) | 11.6 [8.4, 14.8] | 17.4 [13.9, 20.8] |
| ^†^Respondent works in LAX zip code^x,y^ | | | |
| No | 615 (97.5) | 8.3 [6.1, 10.4] | 12.8 [10.3, 15.7] |
| Yes | 16 (2.5) | 31.3 [17.4, 45.1] | 37.5 [20.7, 54.3] |

Notes: Serologic survey conducted in October 2020. ^x^p<0.05 test for difference (manufacturer); ^y^p<0.05 test for difference (modified); ^†^Only N=631 have valid zip code data

In addition to univariate analysis presented in the main paper, multivariable logistic regression was performed (Table 3). The likelihood of seropositivity was most strongly associated with whether the respondent reported that they thought it was likely they had Covid-19 (OR=10.8, 95% CI=5.58-20.9). Being Hispanic was a significant predictor of seropositivity for the manufacturer cutoff (OR=2.39, 95% CI=1.20-4.74), but not the modified cutoff (OR=1.65, 95% CI=0.93-2.93.)

**Table 3. Adjusted odds ratios and 95% CIs for seropositivity based on manufacturer and modified thresholds among firefighters in Los Angeles, CA, USA**

|  | Odds Ratio [95% CI] | |
| --- | --- | --- |
|  | Manufacturer cutoff | Modified cutoff |
| 30-44 years old (ref: 18-29 years) | 1.395  [0.52 – 3.76] | 0.925  [0.44 – 1.95] |
| 45-60 years old | 1.090  [0.39 – 3.05] | 0.753  [0.35 – 1.64] |
| ≥60 years old | 0.902  [0.16 – 5.26] | 0.349  [0.07 – 1.79] |
| Female gender (ref: Male) | 0.490  [0.15 – 1.58] | 0.538  [0.21 – 1.35] |
| Race: Non-Hispanic Black (ref: Non-Hispanic White) | 1.396  (0.417 - 4.675) | 1.817  (0.712 - 4.639) |
| Hispanic | 2.387*  (1.202 - 4.741) | 1.646  (0.926 - 2.928) |
| Non-Hispanic Asian | 2.208  (0.818 - 5.962) | 1.092  (0.433 - 2.758) |
| Other/Mixed Race | 2.567  (0.465 - 14.17) | 4.172*  (1.288 - 13.51) |
| Had a Covid test (ref: no) | 1.074  (0.414 - 2.785) | 1.192  (0.540 - 2.633) |
| Don’t know if had a Covid test | 6.011  (0.189 - 190.9) | 3.389  (0.162 - 70.81) |
| Experienced symptoms since pandemic start (ref: no) | 1.647  (0.880 - 3.082) | 1.374  (0.823 - 2.293) |
| Respondent thought it was likely they had Covid-19 (ref: no) | 10.79*  (5.577 - 20.89) | 8.984*  (5.257 - 15.35) |
| Respondent likely had contact with anyone with known or suspected Covid-19 (ref: no) | 0.899  (0.453 - 1.782) | 1.002  (0.574 - 1.746) |
| Respondent works in LAX zip code (ref: no) | 3.172  (0.856 - 11.75) | 2.601  (0.777 - 8.710) |

Notes: *p<0.05

We analyzed seroprevalence for firefighters’ workplace zip codes based on tertiles of contextual factors (Table 4). Seroprevalence was higher for firefighters who worked in zip codes with higher median income, higher share of insured people, lower unemployment, lower share of people with college degrees, and higher shares of people under 40. The difference in seropositivity across tertiles was only statistically significant (p<0.05) for unemployment using modified cutoffs. Our contextual factor results are robust to excluding firefighters who worked in the LAX zip code (N=16 respondents) who experienced a SARS-CoV-2 outbreak in July 2020.

**Table 4. Association between workplace contextual factors and SARS-Cov-2 seropositivity among firefighters in Los Angeles, California, USA**

|  | % Seropositive [95% CI] | |
| --- | --- | --- |
| Zip code characteristics | Manufacturer cutoff | Modified cutoff |
| Total [N=631] | 8.9 [6.7, 11.1] | 13.5 [11.0, 16.3] |
| Median income | | |
| Bottom tertile (<$47,000) | 7.7 [4.2, 11.2] | 11.3 [7.1, 15.4] |
| Middle tertile | 6.4 [3.0, 9.8] | 11.3 [7.0, 15.7] |
| Top tertile (>$64,000) | 12.6 [8.1, 17.2] | 18.4 [13.2, 23.7] |
| % with health insurance | | |
| Bottom tertile (<70%) | 8.1 [4.5, 11.8] | 13.6 [9.1, 18.1] |
| Middle tertile | 7.0 [3.4, 10.5] | 10.0 [5.8, 14.1] |
| Top tertile (>91%) | 11.5 [7.2, 15.8] | 17.2 [12.1, 22.3] |
| % unemployed ^y^ | | |
| Bottom tertile (<6%) | 12.1 [7.8, 16.4] | 20.2 [14.9, 25.4] |
| Middle tertile | 6.6 [3.7, 9.6] | 10.7 [7.0, 14.4] |
| Top tertile (>7%) | 8.0 [3.5, 12.6] | 8.8 [4.0, 13.5] |
| % with college degree | | |
| Bottom tertile (<27%) | 10.4 [6.3, 14.5] | 15.6 [10.7, 20.4] |
| Middle tertile | 6.7 [3.3, 10.1] | 11.5 [7.2, 15.8] |
| Top tertile (>40%) | 9.5 [5.6, 13.5] | 13.8 [9.1, 18.5] |
| % ≥65 years old | | |
| Bottom tertile (<12%) | 10.4 [6.3, 14.6] | 17.5 [12.4, 22.7] |
| Middle tertile | 7.5 [4.3, 10.6] | 10.8 [7.1, 14.5] |
| Top tertile (>15%) | 9.2 [4.6, 13.8] | 13.2 [7.8, 18.5] |
| % ≤40 years old | | |
| Bottom tertile (<54%) | 9.5 [5.7, 13.3] | 13.4 [9.0, 17.8] |
| Middle tertile | 7.3 [4.2, 10.3] | 12.0 [8.2, 15.8] |
| Top tertile (>59%) | 11.2 [5.7, 16.7] | 17.6 [10.9, 24.3] |
| % white, non-Hispanic | | |
| Bottom tertile (<16%) | 7.4 [4.4, 10.4] | 11.4 [7.8, 15.0] |
| Middle tertile | 7.3 [2.7, 11.8] | 12.9 [7.0, 18.8] |
| Top tertile (>36%) | 12.0 [7.6, 16.4] | 17.2 [12.2, 22.3] |

Notes: ^x^p<0.05 test for difference (manufacturer); ^y^p<0.05 test for difference (modified)
